# Outcomes Of Carotid Artery Stenting Without Embolic Protection in Yemen: A Resource-Constrained Experience

**DOI:** 10.1101/2025.06.18.25329892

**Authors:** Aussan Al-Athwari

## Abstract

**BACKGROUND:** Carotid Artery stenting (CAS) have been reported to be safe and effective option for treating carotid atherosclerotic diseases. The published data support the use of embolic protection devices (EPDs) to reduce periprocedural stroke. However, reports and studies from resource limited countries are scarce. This study aimed to evaluate the outcome of CAS procedures without EPDs in Yemen as one of the lowest-income countries.

**METHODS:** This is a retrospective cohort study regarding carotid artery stenting (CAS) for symptomatic carotid artery stenosis that was conducted at the stroke center of and American Modern Hospital (AMH) during the period from March 2023 to March 2025. All Patients with symptomatic carotid artery stenosis were included in the study. CAS procedures were performed by a single interventional neurologist. The primary outcome included a 30-day periprocedural mortality, stroke, and myocardial infarction or arrythmia. Any other complications are considered secondary outcomes.

**RESULTS:** A total of 62 (53 males) patients included in this study, mean age 60.2 ± 9.68 years. All Patients had symptomatic carotid artery stenosis. Technical success rate was 100%. (%). One patient developed transient dysarthria but DW-MRI was negative. Significant bradycardia occurred in two patients and responded immediately to atropine. Three patients developed mild local hematoma and one patient had femoral pseudoaneurysm. Closed-Cell Carotid Wall Stent (Boston Scientific) was used as a single stent in all patients.

**CONCLUSION:** CAS conducted by a trained interventional neurologist without EPDs, demonstrates a low complication rate and it is effective and safe option in countries with-limited resources.

## INTRODUCTION

Eestablishment of stroke centers have become the standard of care for all stroke patients to reduce death and disability. ^1^ Almost 89 % of these stroke related deaths and disabilities occur in low-middle-income countries (LMIC). ^2^’^3^ Approximately 20% of strokes are caused by Carotid atherosclerosis with only a limited number of studies or clinical trials that explore the outcomes of carotid revascularization for patients with carotid artery diseases in resource-limited countries. ^4^’^5^

Carotid artery stenting (CAS) is a minimally invasive alternative to CEA with comparable outcome.^6^’^7^ CAS is associated with higher embolic complications risk than carotid endarterectomy (CEA) so the embolic protection devices (EPDs) are used to decrease this risk.^8^’^9^’^10^ However, duo to mainly financial concerns, many studies in low-income countries reported no major differences and in experienced hands CAS without EPD can safely be performed. ^11^’^12^’^13^

Yemen is a resource-limited country that has been overwhelmed by devastating political crises and military conflicts since 2011. Providing stroke care services in such conditions is being challenged by infrastructure limitations, training and expertise shortage, high procedural costs and financial constraints, ongoing conflicts and humanitarian crisis, ^14^ so the experience of the first neurointerventional experience in Yemen need to be evaluated.

## SUBJECTS AND METHODS

### Study design and patient Selection

This retrospective observational study explored the outcome of CAS without EPDs for symptomatic extracranial stenosis. It was conducted between March 2023 and March 2025 at the first and only interventional neurology center-American Modern Hospital in Aden city, Yemen. The data of All adult (> 18) patients were included. The degree of stenosis assessed by duplex ultrasound (US) imaging or CT angiography and confirmed by digital subtraction angiography (DSA) and calculated according to formula proposed by NASCET method. ^15^ [**Rothwell et al**.] Carotid stenosis was defined as symptomatic ≥ 60% stenosis. Patients with complete occlusion or incomplete data were excluded. Demographic data and risk factors (diabetes mellitus, hypertension, dyslipidemia and current smocking (or within last year) were screened in all patients. Neurological examination is performed for all patients and investigations included routine labs, duplex, Echocardiography.

### CAS procedure

All CAS procedures were performed by a single interventional neurologist (2 years of training as a subspeciality) and the stents used were all carotid WALLSTENT (Boston Scientific, USA). The patient should be on dual antiplatelets (aspirin 75 mg and clopidogrel 75mg for at least 5 days before the procedure or aspirin 75 and ticagrelor 90 mg twice daily). In patients who were on aspirin only the received ticagrelor loading 190 mg at least 2 hours before the procedure. Dual antiplatelets continued for 6 months after the procedure then switch to a single antiplatelet usually aspirin 100 mg once daily.

The common femoral artery used in all patients. Under local anesthesia a short 10 cm Terumo (Terumo Medical, Tokyo, Japan) 8-French sheath was inserted into the common femoral artery followed immediately by administration of 5,000 international units of heparin. First a 5 F diagnostic catheter Bern (Cordis, USA) used to do imaging of all arteries and then an 8-French guiding-catheter (Mach1 SoftTip, Boston Scientific, USA) is placed in the common carotid artery under continuous heparin infusion. If angioplasty is needed before stenting, then we use 2.5 * 20 mm and we use 5.5* 20 for post-dilatation angioplasty with a goal of covering the lesion and residual stenosis less than 30%. Atropine is kept ready in syringe for any bradycardia. After the stent is deployed, final imaging of vessels to exclude distal embolism and the catheter is removed. Finally, Manual compression is applied to the access site for 20-30 minutes to prevent bleeding and the patient is monitored in ICU overnight for any complications such as bleeding or stroke.

### Outcomes measures

Neurological evaluation was performed using the national institute of health stroke scale (NIHSS) by a well-trained neurologist just before the procedure, immediately after and at 12 hours later before discharge. Complications associated with CAS occurring up to 30 days postoperatively were evaluated. Postoperative Neuroimaging is ordered for patients who develop periprocedural neurological symptoms and on day 30 carotid duplex routinely performed. The CAS procedure is considered successful if the residual stenosis is less than 30 %.

### Statistical Analysis

Data analysis was done using the Statistical Package for Social Sciences (SPSS) version 23.0 (IBM Co., Armonk, NY, USA).

## RESULTS

A total of 62 patients were included in this retrospective study. fifty-three patients were male (85.48%), while nine were female (14.52 %). Mean age of patients was 60.2 ± 9,6 years. All patients treated successfully with deployment of a single closed-cell stent (Carotid wallstent). The site of stent deployment was left ICA in 43 patients (69.35 %) and right MCA in 19 patients (30.65%). NIHSS before procedure was zero in 47%, 3-5 in 34% and 6-9 in 19% of patients. The mean NASCET was 76 ± 10 %. Predilation angioplasty was needed in 16 (25.8 %) patients and postdilation angioplasty in 58 (93.55 %%) patients. Significant bradycardia occurred in two patients and responded immediately to atropine. One patient developed transient dysarthria but DW-MRI was negative. Three patients developed mild local hematoma and one patient had pseudoaneurysm. No stroke, myocardial infraction or in-stent thrombosis documented during the 30-day follow up. A summary of the baseline characteristics and risk factors details is presented in Table 1.

**Table 1.** Characteristics of the studied patients.

| Characteristics |  | Total no. of patients<br>(62) |
| --- | --- | --- |
| Age (mean ± SD) | | 60.2 ± 9.68 |
| Gender | Male | 53(85.48 %) |
|  | Female | 9 (14.52 %) |
| Symptomatic |  | All |
| NASCET (% ± SD) | | 76 ± 10 % |
| Risk Factors (%) |  |  |
| • *DM* |  | 31 (50%) |
| • *Hypertension* |  | 59 (95.16%) |
| • *IHD* |  | 34 (54.84%) |
| • *Dyslipidemia* |  | 57 (91.94%) |
| • *Smocking* |  | 49 (79.03 %) |
| • *Khat chewing* |  | 51 (82.62%) |
| Type of stent |  | Carotid WallStent (100%) |
| EPDs |  | None |

## DISCUSSION

Carotid artery stenting (CAS) has emerged as a minimally invasive alternative to carotid endarterectomy for the treatment of carotid artery stenosis, particularly in patients at high surgical risk. This procedure has gained widespread acceptance in high-income countries with advanced medical infrastructure and in the United States CAS is reimbursed for symptomatic patients with ≥50% carotid stenosis and asymptomatic patients with ≥70% stenosis by the Centers for Medicare and Medicaid Services issued a new National Coverage Determination. ^16^ However, its implementation in limited-resources settings presents unique challenges. These challenges include restricted access to specialized equipment, limited availability of trained interventionalists, and financial constraints that hinder the adoption of advanced medical technologies.14 The use of Embolic Protection Devices (EPDs) and closure systems is presenting extra cost challenge, so the feasibility and outcomes of CAS in resource-limited settings remain underexplored, with limited data available to guide clinical practice.

This study evaluates the implementation, challenges, and outcomes of carotid artery stenting without EPDs in a limited-resources country, focusing on the experience at American Modern Hospital in Aden, Yemen.

The first diagnostic cerebral angiography in Yemen was performed in February 2023 together with the establishment of the first comprehensive stroke center. The commonest procedure performed at the center is the CAS. the overall technical success rate was 100%which compares well with the published data.

There are no big studies about the prevalence of extracranial atherosclerosis in Yemeni patients. In a small study conducted in Dhamar university among 84 patients included in the study carotid stenosis found in 45 patients (54 %) of patients and a significant stenosis (> 60%) was found in in 23 patients (27.38 %). ^17^ The predominant risk factors were smocking, hypertension, dyslipidemia and DM respectively. Khat chewing is included as many studies supports the association of this habit with cardiovascular diseases and premature atherosclerosis which could explain the younger mean age of the population in our study. ^18^’^19^’^20^’^21^’^22^

The neurointerventionalist’s major concern during CAS procedure is the cerebral embolism. To reduce the incidence of periprocedural embolic stroke EPDs are recommended (Class IIa, level B) and there are two types, distal EPDs and proximal EPDs. ^23^’^24^’^25^’^26^’^27^’^28^’^29^’^30^ However, many studies argue the significance of protected CAS. ^31^’^32^’^33^’^34^’^35^ and some believe that the unprotected techniques may decrease neurological sequelae especially in tortuous arteries where EMDs can offer extra manoeuvring that can lead to embolism, dissection or vasospasm. ^36^

The recommendation of EPDs duo to higher stroke in unprotected group such as EVA-3S trial is questioned for the later incidence of stroke and the age of unprotected group was 8 years older. ^37^ The risk of cerebral embolization is present during all stages of CAS: passing the lesion with a wire, pre-dilatation, placement of the protection device, stent deployment, and post dilatation. ^38^’^39^

Closed cell designed stents can provide better scaffolding to the carotid lesion and hence decrease the danger of plaque extrusion via the interstices of the stent during its deployment, post dilatation, and after finishing the procedure. ^40^ Two previous studies showed a trend of better outcome with closed cell stents.^41^’^42^

No studies specifically addressed the cost-effectiveness of CAS in Yemen. However, anecdotal evidence suggested that CAS could be cost-effective in selected patient populations, particularly when compared to the long-term costs of stroke care. This study highlights the potential of CAS as a viable treatment option for carotid artery stenosis in Yemen, despite significant challenges. The clinical outcomes observed in Yemeni patients are comparable to those in other low-resource settings, underscoring the importance of expanding access to this life-saving procedure. However, the success of CAS in Yemen depends on addressing key barriers, including infrastructure limitations, training gaps, and financial constraints.

## CONCLUSION

Carotid artery stenting is a promising intervention for stroke prevention in limited-resources countries, but its widespread adoption requires targeted efforts to overcome systemic challenges. This study underscores the need for further research and investment to ensure equitable access to CAS and improve outcomes for patients with carotid artery stenosis.

## Data Availability

Data available upon request

## Abbreviations

CAS: Carotid Artery Stenting
CEA: Carotid Endarterectomy
EPDs: Embolic Protection Devices
DW-MRI: diffusion-weighted magnetic resonance imaging
NIHSS: national institute of health stroke scale
NASCET: North American Symptomatic Carotid Endarterectomy trial
LMIC: low-middle-income countries

